# NIR autofluorescence allows for pituitary gland detection during surgery: the first evidence from microscopic studies and in vivo measurements

**DOI:** 10.64898/2026.03.05.26347733

**Authors:** Veronika Alibaeva, Nadezhda Korneva, Andrey Grigoriev, Grigoriy Starkov, Gleb Budylin, Vilen Azizyan, Anastasia Lapshina, Nano Pachuashvili, Ekaterina Troshina, Natalia Mokrysheva, Liliya Urusova, Evgeny Shirshin

## Abstract

A critical challenge in endocrine neurosurgery is intraoperative discrimination between normal pituitary tissue and pituitary neuroendocrine tumors (PitNETs). Suggesting the universal persistence of near-infrared autofluorescence (NIRAF) in endocrine organs and inspired by routine clinical use of NIRAF for parathyroid gland identification, we discovered that pituitary NIRAF can be employed for label-free transsphenoidal surgery guidance. *Ex vivo* confocal spectral imaging of 33 specimens identified secretory granules as the dominant long-wavelength fluorescence source and showed that normal pituitary had higher granule content than PitNETs. For the first time, we made use of the pituitary NIRAF during surgery and assessed its performance for pituitary/adenoma separation *in vivo* for 27 surgeries and showed near-perfect separability between pituitary and non-pituitary measurement sites with ROC-AUC of 0.98. The obtained results clearly demonstrate that the suggested method, based on the solid microscopic background, has the potential for clinical translation and paves the way for enhanced gland preservation during resection.

## 1. Introduction

Pituitary neuroendocrine tumors (PitNETs) account for ∼15% of all primary brain tumors and are the third most common intracranial neoplasm [1]. PitNETs are treated surgically when they reach clinically significant sizes or persist in hormonally active forms. With the exception of prolactinomas, which typically respond to medical therapy, endoscopic endonasal resection remains the standard initial treatment for symptomatic disease and usually yields clinical improvement [2]. Nevertheless, gross-total resection is achieved in only about 80% of cases, and regrowth from residual tumor has been reported in up to 75% after incomplete removal [3]. A key contributor to suboptimal outcomes is the difficulty in distinguishing PitNETs from normal pituitary intraoperatively, especially in fibrotic or infiltrative tumors [4]. Moreover, inadvertent damage of normal pituitary tissue during surgery can lead to endocrine disorders. Hence, new methods for intraoperative discrimination between pituitary, tumor and surrounding tissues are of high demand.

Various exogenous fluorescent agents were tested for intraoperative visualization of PitNETs, including chlorin e6 (Ce6), 5-aminolevulinic acid (5-ALA), indocyanine green (ICG), OTL38, fluorescein, and labeled antibodies (e.g., bevacizumab-800CW). Each agent has a distinct accumulation mechanism, dosing/timing protocol, and tumor-to-normal contrast profile. Among these, ICG has the most clinical experience to date [5–7]. Normal pituitary and adenomas are discriminated by the relative ICG fluorescence intensity at certain times after injection, and numerous studies aim at the selection of proper time window and injection protocol (e.g., using the second window [8]) for tumor margin delineation [9–13]. Despite the increase of positive outcomes when using ICG guidance, some works emphasize sensitivity of this method to artifacts: strong background signal from sphenoid sinus mucosa and blood clots in the field can obscure or suppress the NIR ICG signal [7,14]. Chlorin e6 (Ce6) was tested as optical contrast in patients with pituitary macroadenomas, and a high concentration of Ce6 was observed in the tumor tissue: after resection no foci of increased fluorescence and no residual tumor areas were detected, indicating effective visualization of the adenoma outlines. However, there is a difficulty in interpreting the images and the need to optimize the administration time and dose for different sizes of adenomas [15]. Another optical contrast agent, 5-ALA, is metabolized to PpIX and provides intraoperative fluorescence of tumor tissue [16]. 5-ALA is widely used for fluorescence guidance in neurosurgery, employing both steady-state and time-resolved detection methods [17–19]. It has been also tested for contrasting PitNETs, while the results were somewhat contradictory [20–22].

Targeted optical contrast agents include the folate-targeted probe OTL38, which delivers strong near-infrared fluorescence in nonfunctioning pituitary adenomas with high folate receptor alpha (FRα) expression, supporting subtype-selective visualization [23]. In contrast, in cases of low FRα expression, the signal-to-background ratio (SBR) declines significantly. The VEGF-targeted agent bevacizumab-800CW produces stable fluorescence near 800 nm with a dose-dependent increase in signal. Yet, VEGF expression did not correlate with PitNET subtype, limiting predictive specificity [24]. Collectively, these studies indicate that while targeted and vascular probes can yield intraoperative contrast under optimized conditions, their performance remains constrained by variable expression profiles, nonspecific uptake, and protocol complexity. Hence, the development of label-free methods for fluorescence surgery guidance is highly desirable.

In the recent decade, it has been shown that endocrine tissues have a pronounced NIR autofluorescence (NIRAF), suitable for intraoperative navigation. For instance, the PTeye probe system based on parathyroid gland (PTG) NIRAF was implemented in clinical practice. In a multicenter randomized study, the use of PTeye during thyroidectomy and bilateral cervical revision significantly increased the number of confidently identified parathyroid glands without prolonging surgery [25]. Ex vivo studies of resected adrenalectomy specimens showed pronounced NIRAF in normal adrenal tissue. Adrenal tumors also exhibited NIRAF [26], but with pathology-dependent intensity: cortisol-producing tumors showed the highest signal, whereas some other tumor types had weaker fluorescence.

Suggesting the universal persistence of NIRAF in endocrine organs, we questioned whether the pituitary gland may have intrinsic autofluorescence sufficient for surgery guidance. Starting from verification of this hypothesis, we performed fluorescence imaging of normal pituitary and PitNETs at macro and micro levels with confocal microscopy. The comprehensive analysis of fluorescence localization upon different excitation regimes, including NIRAF, allowed for determination of optical differences between pituitary and tumors, which were used as the basis for the development of the intraoperative navigation device. Finally, for the first time, we made use of the pituitary NIRAF during surgery and assessed its performance for pituitary/adenoma separation *in vivo*. The obtained results clearly demonstrate that the suggested method, based on the solid microscopic background, has the potential for clinical translation and paves the way for enhanced gland preservation during resection.

## 2. Materials and methods

### 2.1 Ex vivo samples preparation

All *ex vivo* tissue specimens were collected at the National Medical Research Center of Endocrinology named after Academician I.I. Dedov, Moscow, Russia from people with planned surgical interventions, with written consent. The study was approved by the local ethics committee under protocol No. 17 dated 10 September 2025.

For macroscopic NIRAF imaging, ∼1×1×1 cm^3^ tissue samples of the normal thyroid, parathyroid adenoma, adipose tissue adjacent to thyroid, pituitary and adrenal glands from various patients were used, obtained on the day of surgery within no more than 2-3 hours after surgery.

For confocal imaging, fresh postoperative human pituitary surgical specimens were collected. In total, 40 specimens were included and classified as 22 somatotroph PitNETs, 11 corticotroph PitNETs, 4 clinically non-functioning PitNETs, 2 prolactinomas, and 1 thyrotroph PitNET. 9 specimens that were initially collected as presumed tumor tissue sites but were subsequently identified as normal pituitary tissue on histology were analyzed as a separate normal group. In most cases, one tissue fragment per patient was first imaged by confocal microscopy and then processed for routine histopathology and immunohistochemistry together with the remaining surgical material.

### 2.2 Endocrine macroscopic NIRAF imaging

Macroscopic images were acquired for the same set of *ex vivo* endocrine tissue samples under white light at two excitation wavelengths, 690 nm and 785 nm, while recording the emitted signal in 820–870 nm range with the Mobile Fluorescence Imaging Device (Biomedical Fluorescence Imaging Systems, LLC, Nizhniy Novgorod, Russia) macro-imaging system providing spatial resolution of ∼100 μm/pixel. Autofluorescence images (under 690 nm and 785 nm excitation with intensity ∼10 mW/cm^2^ at tissue surface) were processed using an identical brightness and contrast mapping and a median filter with 2-pixel window.

To quantify tissue fluorescence, one fixed-size circular region of interest (ROI) with an area of 1597 pixels was placed on each tissue sample, thyroid gland, parathyroid gland, pituitary gland, adrenal gland, and adjacent adipose tissue. For each excitation condition, the mean pixel intensity within the corresponding ROI was calculated. The paired mean values from the 690 nm and 785 nm excitation images were subsequently used for correlation analysis between the two excitation regimes.

### 2.3 Pituitary confocal microscopy imaging

All confocal measurements were performed on an Olympus FV3000 (Olympus, Japan) confocal laser scanning microscope using a 60× objective and four excitation wavelengths (405, 488, 561 and 640 nm). Tissue fragments typically ranged from approximately 1 mm to 1 cm in size, were placed in glass-bottom confocal dishes and gently covered with sterile saline buffer to prevent drying. Samples were stored at 4 °C and imaged within 1–6 h after surgical resection at room temperature. Images were acquired as 512×512-pixel fields of view (FOV) with a pixel size of 0.414 µm. For each specimen, approximately 10-15 FOVs were recorded at different locations within the same fragment to capture intra-sample heterogeneity. For comparison with hematoxylin and eosin staining (H&E) larger tissue areas were additionally measured by stitching 4×3 adjacent fields into a single mosaic while maintaining cellular-level resolution.

Morphological imaging was performed in the confocal reflectance mode using a 640 nm laser with narrow-band detection of reflected light at 635-645 nm. To avoid detector saturation, reflectance imaging was carried out with reduced excitation power and lowered detector voltage. Autofluorescence was recorded in spectral (lambda scan) imaging mode with a 10-nm step: 420–800 nm range under 405 nm excitation and 660-800 nm range under 640 nm excitation. Spectrally encoded fluorescence maps were generated by assigning each pixel a representative emission wavelength, computed as the intensity-weighted mean of the pixel’s background-corrected emission spectrum. Pixels with negligible integrated signal were treated as background and displayed with zero brightness. To enable direct visual comparison across samples, the colormap limits were fixed for each excitation wavelength.

### 2.4 Confocal image analysis

To quantitatively analyze confocal images, three classes of regions of interest (ROIs) corresponding to the dominant microscopic contributors to autofluorescence were introduced: granule ROIs, collagen ROIs, and the remaining cellular ROIs, hereafter referred to as cytoplasm ROIs.

Granule ROIs were segmented using the following procedure. First, for each FOV acquired in autofluorescence mode with 640 nm excitation, images were summed along the spectral axis to obtain an integral intensity image. Then, an intensity threshold corresponding to the 90^th^ percentile of the intensity distribution was applied to generate binary mask highlighting bright spots within FOV. To suppress noise and extract morphologically significant regions corresponding to granules – rather than isolated bright pixels – a connected component labeling algorithm with 3x3-kernel was used. Labeled regions with an area lower than 40 pixels (∼7 µm^2^) were filtered out. The remaining segmented ROIs were subsequently used for the analysis, including the calculation of emission spectra and the total area occupied by granules.

Collagen ROIs were identified in datasets using FOVs acquired in autofluorescence mode with 405 nm excitation using a composite criterion that combined spectral similarity to a reference collagen emission spectra and integral intensity mask. As a first step, the “reference” collagen emission spectrum was calculated for manually selected ROI in image with clearly visible collagen fibers as the spectrum averaged across all pixels in the ROI. Next, cosine similarity was calculated between the reference collagen spectrum and spectra in FOVs pixels. Regions with high cosine similarity (above 0.9) and intensities in the range from the 30th to the 95th percentile of the intensity distribution were then selected. Morphological opening and closing operations were then performed with a 3x3 pixel kernel to eliminate artifacts. Cytoplasm ROIs were defined as remaining pixels not assigned to granule or collagen ROIs and exhibiting characteristic cellular structure.

The areas occupied by specific ROI types were determined as follows. First, the area occupied by a specific ROI type was calculated for each FOV. The median area was then calculated for each tissue sample. These median areas were compared between different subgroups (normal pituitary tissue and PitNET subtypes). Similar aggregation procedure was used to obtain the median spectra of the granules. First, the median spectra per ROI for each FOV were obtained, then the median spectra for each sample were obtained among these spectra. As a result, we obtained one representative spectrum for each sample. This scheme ensures the independence of observations and prevents the dominance of one pituitary gland in a large number of frames, while maintaining resistance to outliers by using the median in the group stage. All diagrams and spectra were calculated for 19 samples of the somatotroph PitNETs with 101 FOV, 5 samples of the corticotroph PitNETs with 37 FOV, and 9 samples of the normal pituitary gland with 52 FOV. Per sample median area of collagen and granules ROI were compared between groups using a nonparametric two–sided Mann-Whitney test. P-values were adjusted for multiple comparisons using the Holm-Bonferroni method.

### 2.5 In vivo NIR-autofluorescence measurement setup

NIRAF was recorded using a fiber-optic system with a 650-nm laser source (OXlaser, China), provided excitation light, which was spectrally “cleaned” by a bandpass filter OFBH110-650 (JCOPTIX, China), attenuated using aperture and delivered to the surgical field through a sterile surgical optical fiber. The same fiber collected the emitted signal, which was routed through a two-lens collection system and an 800-nm long-pass filter, OFE1LP-800 (JCOPTIX, China) from the previous version of the system for parathyroid detection [27], to suppress residual excitation light. Spectra were acquired using a spectrometer (YIXIST, China) in 300-1100 nm range with a spectral resolution of 12 nm, and an integration time of 500 ms.

During the study, fibers with NA = 0.22 and different core diameters – 500 µm, 365 µm and 200 µm – were used to improve maneuverability in narrow and curved corridors. The optical power at the fiber tip was maintained within 350-400 µW for all fiber diameters. To account for signal differences between fibers with different core diameters, reference measurements with 1,4 μM methylene blue solution were acquired when switching fibers and were used for calibration of the recorded intensities. Spectrometer background noise was collected with excitation source turned off and was subtracted prior measurements.

### 2.6 In vivo NIR-autofluorescence measurements procedure

The intraoperative *in vivo* study included 27 patients undergoing standard endoscopic endonasal transsphenoidal surgery for pituitary neuroendocrine tumors of which 19 are somatotropinoma, 5 are corticotropinoma, 1 is thyrotropinoma and 5 are hormone-inactive tumors. Optical measurements were integrated into the routine workflow without altering the planned surgical strategy and was approved under the same ethics approval as the *ex vivo* study.

The sterile fiber probe of the setup (Materials and Methods, Section 2.5) was introduced into the surgical corridor under endoscopic guidance and placed in a gentle contact with the target tissue surface. The operating surgeon reported the tissue type at each measurement site (“normal pituitary”, “tumor”, and “other tissues”). These assignments were subsequently confirmed or revised based on postoperative histopathology. The “other tissues” category included dura mater (over the pituitary gland and over the tumor), blood/background, the post-resection tumor bed, coagulated tissue, sphenoid sinus structures, bone, and other non-gland tissues encountered within the surgical corridor.

For quantitative analysis, 5-15 spectra were acquired per measurement site and averaged to reduce noise. From 1 to 6 measurement sites for one or multiple tissue types were measured during surgery. The median NIRAF intensity was computed per each surgery for each tissue category. Within each surgery, all measurement points assigned to a given group were pooled, and a single per surgery median value was calculated. Thus, each surgery contributed at most one value per group, preventing overweighting of surgeries with a larger number of measurements in a given tissue category. Distributions of these surgery-level medians were visualized as boxplots across the three groups. Aggregated median NIRAF values of normal pituitary tissue were compared with tumor and with other tissues using t-test for independent samples. P-values were adjusted for multiple comparisons using Holm-Bonferroni method.

## 3. Results

### 3.1 Endocrine NIRAF

A key practical question for label-free endocrine autofluorescence imaging is the selection of excitation and detection bands. In intraoperative probe measurements performed at 785 nm excitation, parathyroid tissue showed a strong intrinsic fluorescence signal with a peak near 822 nm and higher intensity than adjacent thyroid tissue [28]. Earlier near-infrared autofluorescence spectroscopy using 795 nm excitation reported an emission maximum at 800 nm range for parathyroid tissue [29]. These observations motivate the widely adopted convention of using excitation in the 750–800 nm range and collecting emission in a longer-wavelength window above 800 nm in order to separate fluorescence from excitation leakage and to maximize contrast [30].

Yet, excitation does not need to be restricted to the near-780 nm band. A technical study summarizing endocrine autofluorescence instrumentation discusses both the commonly used excitation range (740–805 nm) with detection typically spanning 800–900 nm and reported examples where the intrinsic signal was elicited using red excitation around 650–685 nm with fluorescence collected above 700 nm [31]. The 650 nm excitation was successfully used for *in vivo* PTG detection in our recent work [27]. In addition, an *ex vivo* multi-wavelength imaging setup explicitly compared visible-red excitation at 628 nm with 780 nm excitation and showed detectable endocrine autofluorescence in both regimes [32].

Motivated by these reports, we performed a direct macroscopic comparison of endocrine autofluorescence imaging under two excitation regimes. For the same set of endocrine organs (thyroid, parathyroid, pituitary, adrenal glands) and adjacent adipose tissue, we acquired images under white light and under excitation at 785 nm and 690 nm at the same excitation intensity of ∼10 mW/cm^2^ while observing the emitted signal in the near-infrared range (Fig. 1a–c). In both excitation conditions, all endocrine tissues produced a clearly detectable autofluorescence signal, whereas adipose tissue remained dim (Fig. 1d). The obtained results clearly demonstrate the pituitary gland exhibits NIRAF close to that of PTG, exceeding that of fat and thyroid gland. Given that the intrinsically high NIRAF of PTG is routinely used for its discrimination from the surrounding tissues during surgery, we hypothesized that the comparably high NIRAF from pituitary can be employed for its detection as well.

**Fig. 1.**
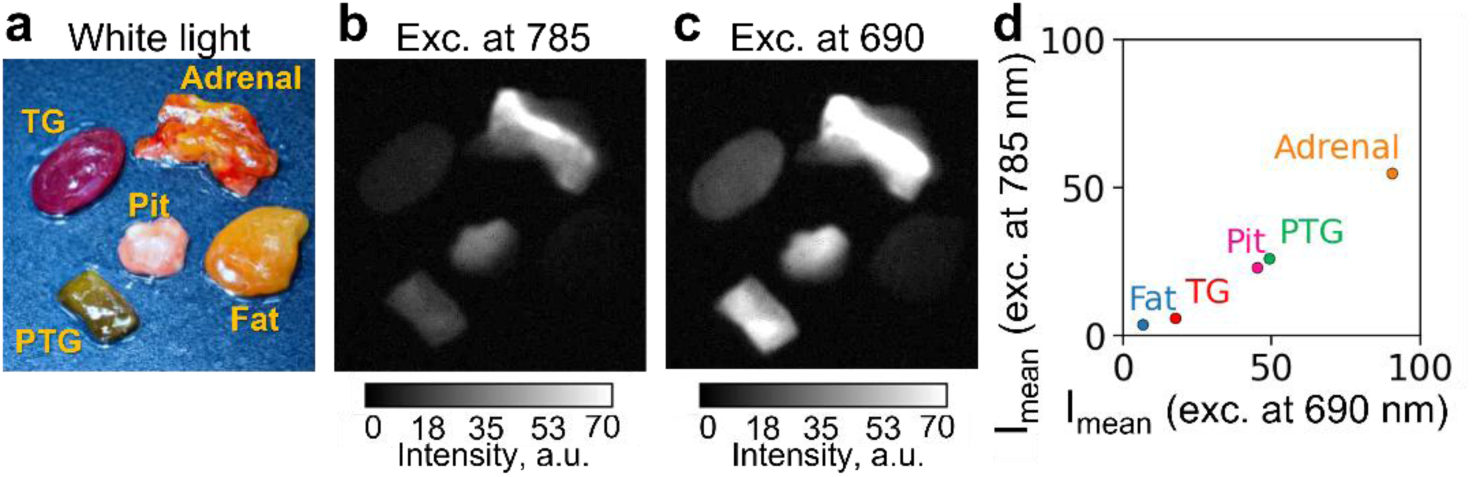
White-light photograph (a) and wide-field near-infrared autofluorescence images acquired under 785 nm excitation (b) and 690 nm excitation (c) for the same *ex vivo* panel of endocrine tissues, thyroid gland (TG), parathyroid gland (PTG), pituitary gland (Pit), adrenal gland (Adrenal), and surrounding adipose tissue (Fat). (d) ROI-averaged autofluorescence intensities measured for each tissue under 690 nm and 785 nm excitation.

We also note that as 690 nm excitation yielded similar NIRAF trends in the adrenal > pituitary ∼ PTG > thyroid ∼ fat sequence, we further used red excitation (640 nm for confocal imaging and 650 nm for *in vivo* detection) to assess the microscopic determinants of elevated NIRAF of pituitary and intraoperative measurements.

### 3.2 Confocal fluorescence imaging of pituitary gland and PitNETs

To assess localization and spectral properties of fluorophores, which may be responsible for NIRAF, in pituitary gland and PitNETs, we performed confocal fluorescence measurements with spectral detection and confocal reflectance imaging, allowing for the analysis of tissue morphology and comparison to histology. In Fig. 2 representative H&E scans (a, d, g), confocal reflectance images (b, e, h) and spectral fluorescence images with excitation at 405 nm (c, f, j) for normal pituitary tissue, somatotroph PitNET and corticotroph PitNET are presented. Normal pituitary tissue showed higher density of bright granules, which were characterized by the red shift of the emission maximum compared to collagen fibers, and a more pronounced fibrous stromal network (Fig. 2c), whereas somatotroph and corticotroph PitNETs (Fig. 2f and j) exhibited a marked decrease in both granular structures and stromal elements. These patterns were consistently observed across the measured samples.

**Fig. 2.**
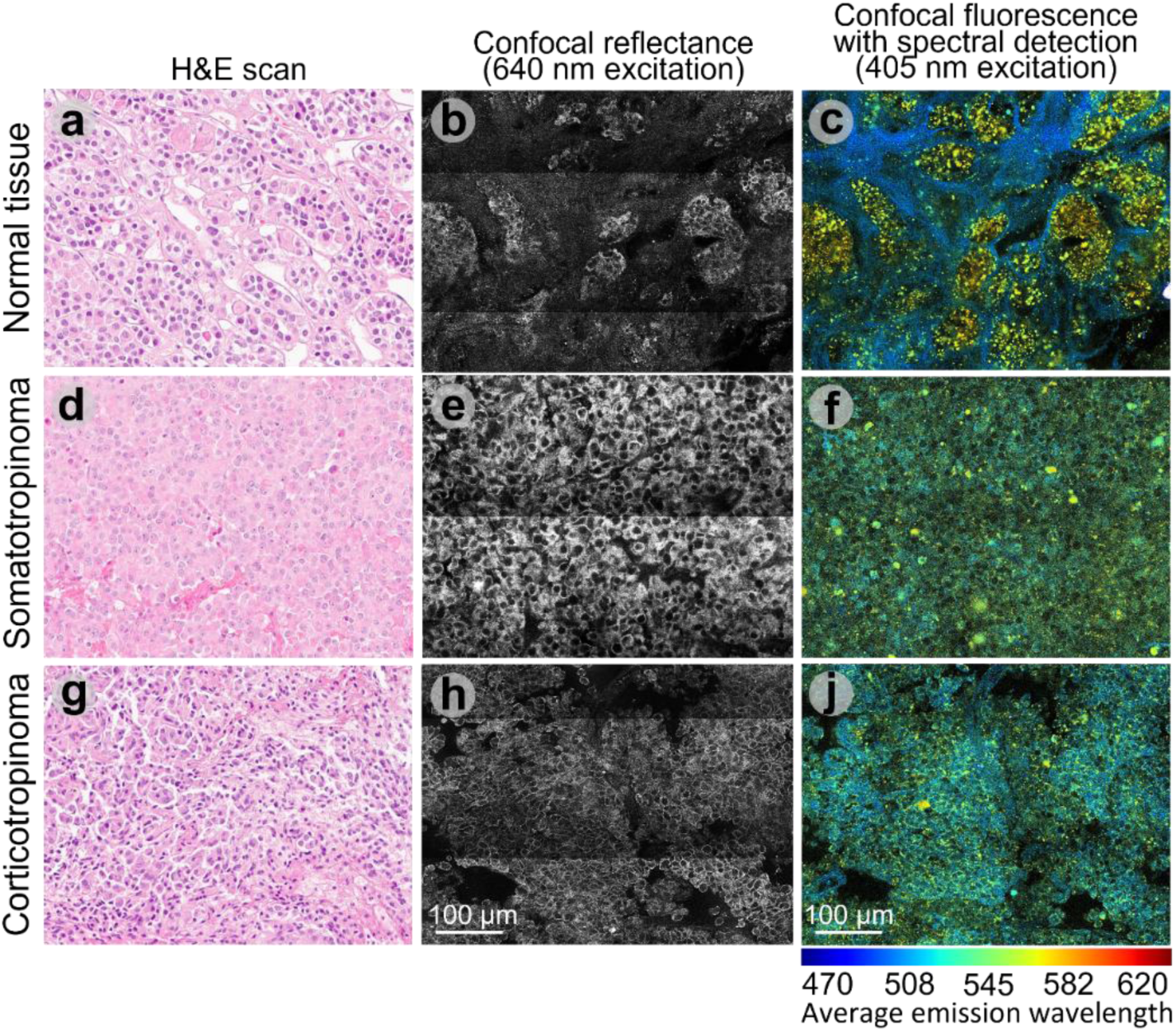
H&E scans (a, d, g), confocal reflectance images (b, e, h), and spectrally encoded confocal fluorescence images acquired under 405 nm excitation (c, f, j) for representative samples of normal pituitary tissue, somatotroph PitNET, and corticotroph PitNET. In the spectral maps, color encodes the weighted-average emission wavelength.

As the fluorescence images of normal pituitary at 405 nm excitation were dominated by intense signal from granules, the lower signal from cells was masked, and individual cells were poorly discernible, in contrast to tumor tissues where a more homogeneous cellular organization became apparent. Confocal reflectance imaging provided complementary structural information by delineating cellular architecture, demonstrating that the bright granules were localized within pituitary cells. Consistent with this, H&E sections of normal pituitary showed a clustered organization of cells separated by collagen-rich fibrovascular stroma, whereas somatotroph and corticotroph PitNETs more often displayed a diffuse cellular growth pattern with reduced intervening stromal framework (Fig. 2b, e, h). The same shift from cell clusters embedded in a fibrous network to a more solid, crowded cellular architecture was observed in the confocal images and motivated the ROI-based quantitative analysis of optical properties.

Across samples, confocal imaging revealed three main ROI classes contributing to autofluorescence: collagen, cytoplasm, and granules (Fig. 3a). The fluorescence spectra extracted from these ROIs under 405 nm excitation showed distinct spectral profiles (Fig. 3b). Secretory granule ROIs exhibited the most red-shifted spectra with maximum at ∼550 nm and a pronounced long-wavelength tail extending up to 800 nm, collagen ROIs showed the most blue-shifted spectra with maximum at ∼470 nm, and cytoplasm ROIs had an intermediate spectral shape. Under 405 nm excitation the total collagen area per FOV was consistently higher in the normal group, whereas somatotroph and corticotroph PitNETs exhibited reduced collagen ROI area (Fig. 3c). This difference was significant by a two-sided Mann–Whitney U test on specimen-level medians with Holm correction for the two planned comparisons: normal versus somatotroph and normal versus corticotroph (both p-values < 10^-3^).

**Fig. 3.**
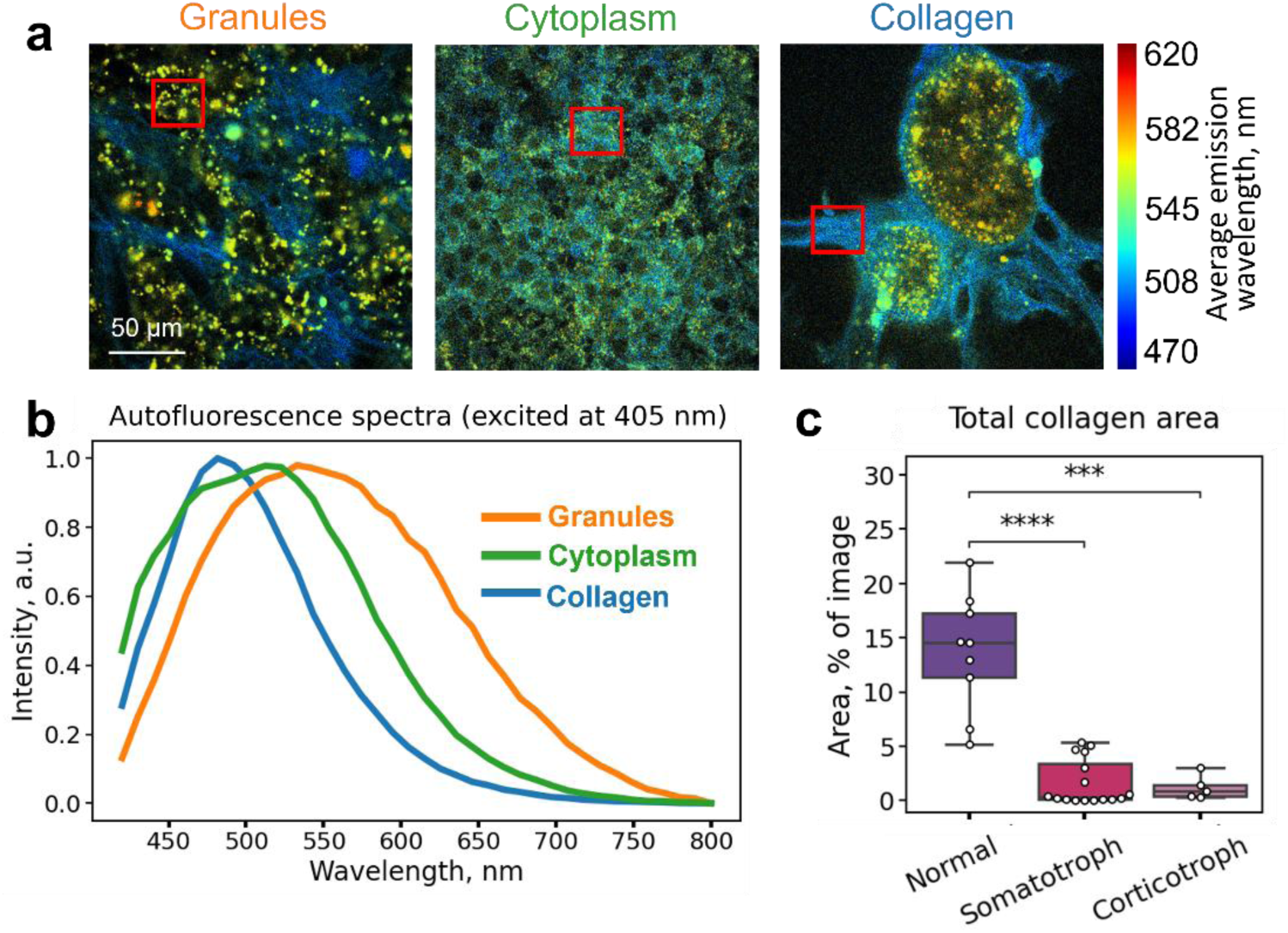
(a) Representative fluorescence spectral images (colors correspond to the intensity-weighted mean emission wavelength) illustrating ROIs dominated by collagen-rich connective tissue, cellular cytoplasm, and secretory granules. (b) Normalized autofluorescence emission spectra extracted from the corresponding ROIs under 405 nm excitation. (c) Boxplots of the collagen ROI area fraction per sample for each tissue type.

We also analyzed fluorescence of normal pituitary and PitNETs under 640 nm excitation to reveal morphological structures responsible for the red/NIR autofluorescence (Fig. 4). Figures 4a-c show the same FOV fluorescence integral intensity excited at 405 nm (Fig. 4a), at 640 nm (Fig. 4b), and as an overlay of the two channels, respectively (Fig. 4c). It was observed that fluorescence, excited at 640 nm, is localized within the granules (Fig. 4b). Moreover, the same granules are excited at 405 nm excitation (Fig. 4a), that is evidenced from the colocalization of signals obtained at 405 nm (blue) and 640 nm (red) excitation – the merged signal provided violet color (Fig. 4c). In contrast, collagen-rich fiber-like structures remain predominantly blue, indicating strong signal under 405 nm excitation and absent signal under 640 nm excitation.

**Fig. 4.**
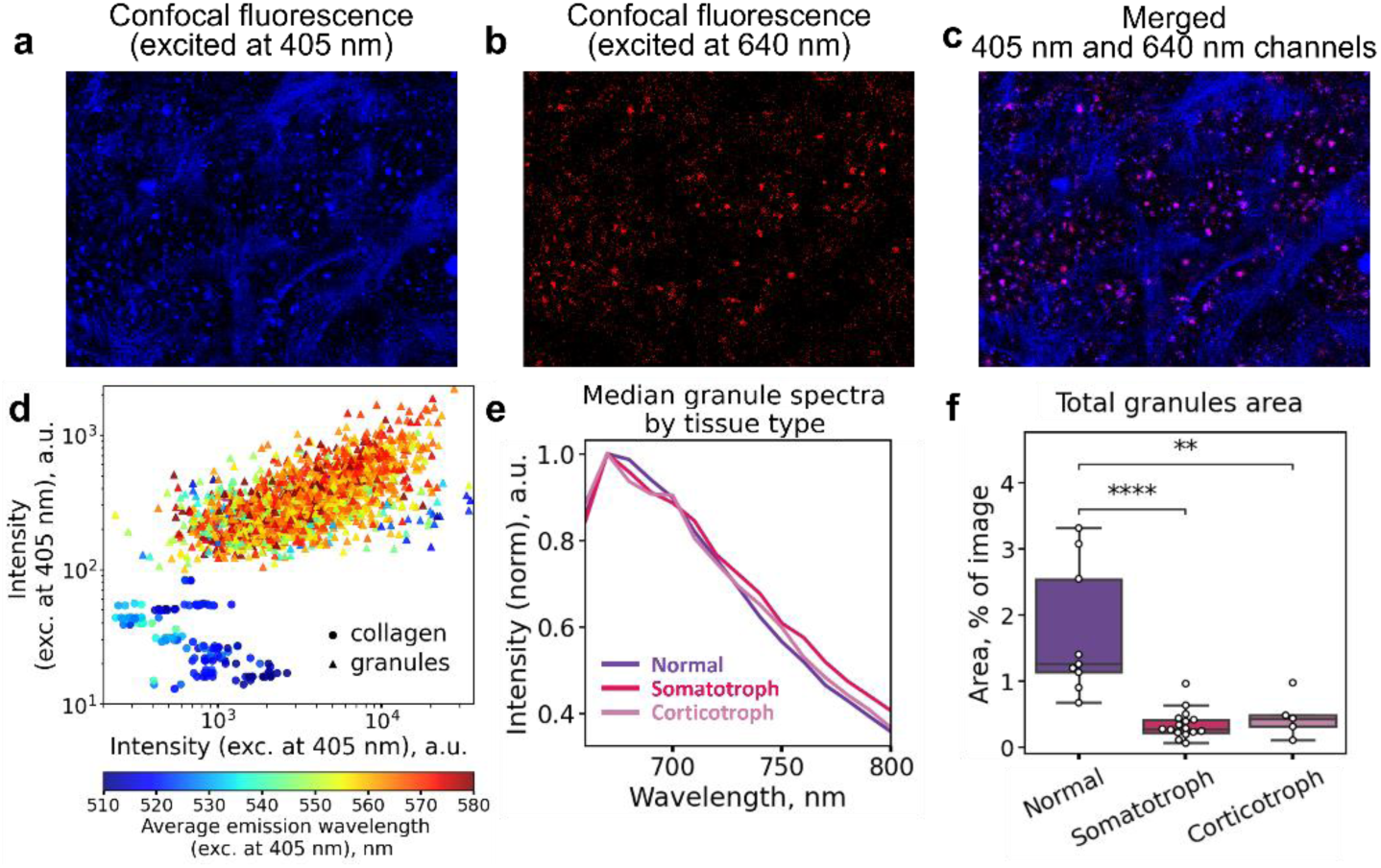
(a–c) Representative confocal spectral images of the same FOV acquired at 405 nm excitation, at 640 nm excitation, and their overlay (blue, red, and magenta co-localization correspondingly). (d) Scatter plot of granules (triangle) and collagen (circle) ROIs autofluorescence intensity under 640 nm versus 405 nm excitation, colored by the intensity-weighted emission wavelength under 405 nm excitation. (e) Normalized median granule emission spectra by tissue type under 640 nm excitation. (f) Boxplots of total granule ROI area expressed as a percentage of image area for each tissue type.

Fig. 4d demonstrates scatter plot of ROI-level autofluorescence intensities measured under 405 nm and 640 nm excitation for 15 random FOVs of all normal pituitary samples. Each point represents one ROI from collagen or granule masks. Granule-associated regions populate an extended high-intensity ridge with simultaneously elevated 405 nm and 640 nm peaks, forming a broadened diagonal band, which supports the colocalized violet granules in Fig. 4c. Collagen-dominated regions concentrate near the bottom of the plot with persistently low 640 nm intensity, consistent with selective excitation at 405 nm. Together, this intensity–intensity analysis provides a quantitative counterpart to the overlay by separating granules and collagen, based on their excitation-dependent fluorescence signatures.

Hence, the bright granules with a red shifted spectrum (maximum at 550 nm, Fig. 3b) observed for the normal pituitary and PitNETs (yellow regions in Fig. 2) can be excited in a broad spectral range, at least from 405 to 640 nm. Finally, the normalized spectra of the granules responsible for fluorescence at 640 nm excitation for normal tissue, somatotroph PitNET, and corticotroph PitNET groups exhibited similar band shape (Fig. 4e), suggesting similarity of the emission source. To assess granule-related differences between diagnostic groups, we quantitatively analyzed the fraction of the image area occupied by the granule ROI, expressed as a percentage of the FOV (Fig. 4f), with normal pituitary exhibiting a larger granule area fraction than both tumor subtypes (*p* < 10^-3^).

Taken together, these results, obtained on a microscopic level, provide quantitative evidence that normal pituitary tissue differs from somatotroph and corticotroph PitNETs by higher abundance of bright granule-associated microdomains and larger collagen-rich stromal component. Given the facts that pituitary exhibits NIRAF at >640 nm excitation, this fluorescence originates from the granules within pituitary cells, which exhibit broad excitation spectrum, and the abundance of granules (Fig. 4f) is elevated in normal pituitary compared to PitNETs, we decided to verify whether pituitary can be identified during surgery using its NIRAF signal.

### 3.3 NIRAF of pituitary gland during surgery

The intraoperative *in vivo* study included 27 patients undergoing standard endoscopic endonasal transsphenoidal surgery for pituitary neuroendocrine tumors. As the initial data, surgeons operate with preoperative MRI, which provides anatomical context and indicates the approximate spatial relationship between the gland, tumor, and surrounding structures. Representative MRI scan for a patient, for whom intraoperative NIRAF measurements were performed, and the zoomed images of the sellar region, as well as the location of the pituitary gland, tumor and dura in it are shown in Fig. 5a. The MRI data is used by surgeons to plan the operation strategy and provides general information about tumor localization, while all the manipulations are performed under endoscopic visualization. However, the tumor and the healthy pituitary gland are still nearby and their boundary may be blurry. The schematic of intraoperative measurements is presented in Fig. 5b. During the surgery, the surgeon can choose the type of nozzle - curved or straight for easier access to tissues, due to the fact that the diameter of the fiber allows for lossless measurements.

**Fig. 5.**
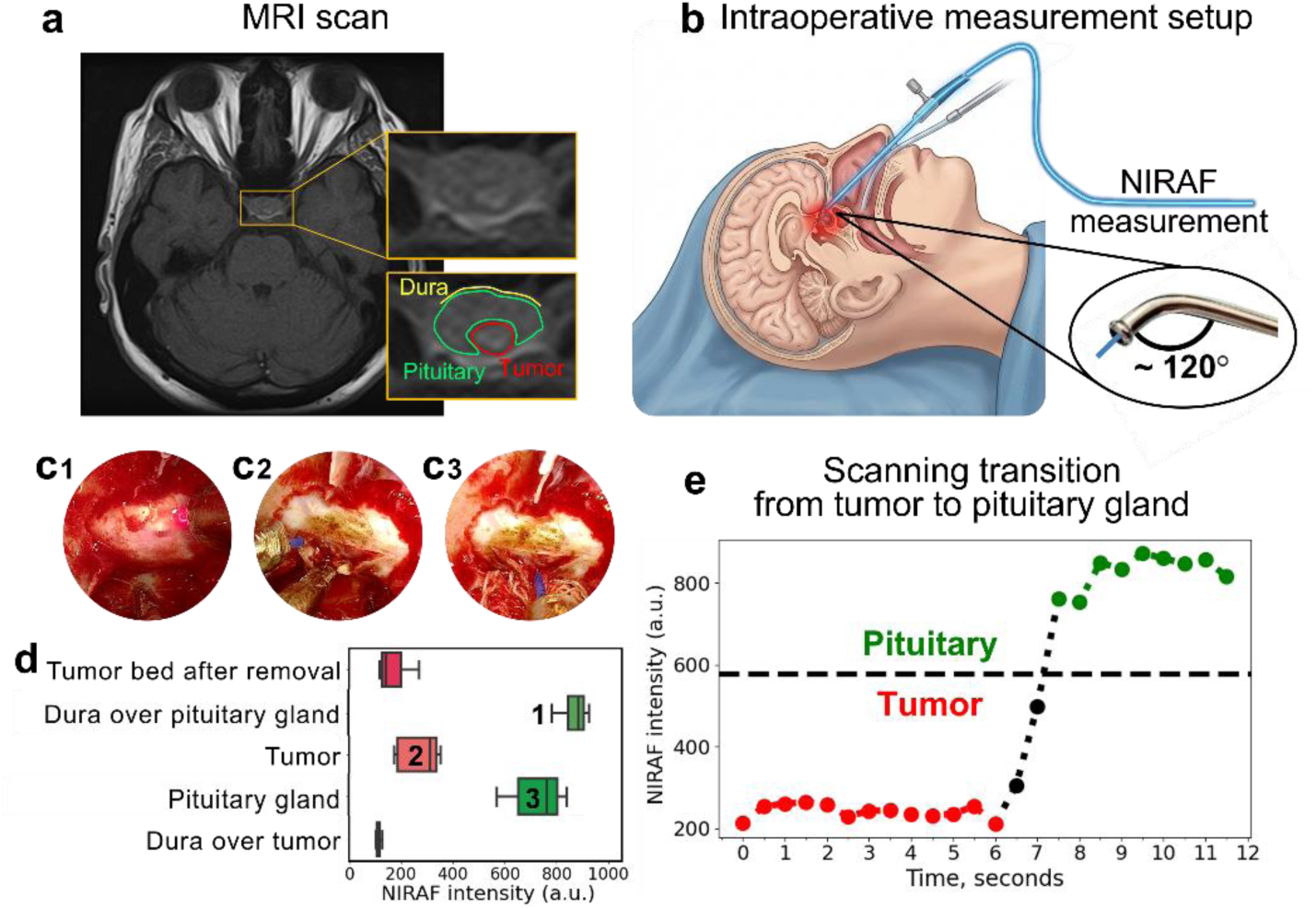
(a) Preoperative MRI for the same patient with a region of interest (sellar region) indicated with zoomed-in view and schematic delineation of the pituitary gland, tumor and dura (b) Schematic of the endoscopic endonasal transsphenoidal access and introduction of the optical fiber to the surgery region. (c1-c3) Endoscopic white-light views showing fiber-probe contact with (c1) dura over the pituitary gland, (c2) tumor, and (c3) normal pituitary gland. (d) Boxplot of NIRAF intensity measured during the same surgery for five tissue classes. (e) Representative time trace showing the transition from the tumor and then onto normal pituitary tissue.

White-light endoscopic views in Fig. 5c (1-3) show the fiber probe in contact with visually identified tissue sites, including the pituitary gland, tumor surface, and dura over the pituitary gland before the incision. Representative NIRAF intensities of different tissue types obtained for the same one surgery are demonstrated in Fig. 5d, it can be seen that NIRAF intensity was highest at the pituitary gland compared with the adjacent non-pituitary sites, demonstrating a clear contrast between pituitary and surrounding tissues under routine surgical conditions.

To demonstrate the kinetics of the NIRAF signal during continuous movement of the probe, we performed measurements while shifting the fiber probe along adjacent structures within the same surgical corridor. Fig. 5e shows a representative time trace recorded during one continuous pass, with consecutive segments corresponding to probe contact with tumor, and visually normal pituitary tissue. The dashed line indicates an empirical contrast level (578 a.u.), computed across all surgeries with measurements in both categories as the median of per-surgery differences between the pituitary and the non-pituitary median NIRAF intensity. The trace highlights plateau-like intervals associated with each tissue type and step-like transitions that occur when the probe crosses tissue boundaries.

Finally, we aggregated measurements from all 27 surgeries and compared NIRAF intensity across tissue classes (Fig. 6a). The boxplots summarize the distributions for the pituitary gland, tumor and other tissue measurement locations. Pituitary sites yielded the highest NIRAF values. As the “other tissues” class includes heterogeneous sites (e.g., dura, blood/background, and post-resection surfaces), partial overlap and occasional intermediate values are expected, particularly for measurements acquired at tissue interfaces. The pituitary gland showed higher median intensities than the non-pituitary tissue classes (Fig. 6a): pituitary gland vs tumor (*p* < 10^-3^) and pituitary gland vs other tissues (*p* < 10^-3^).

**Fig. 6.**
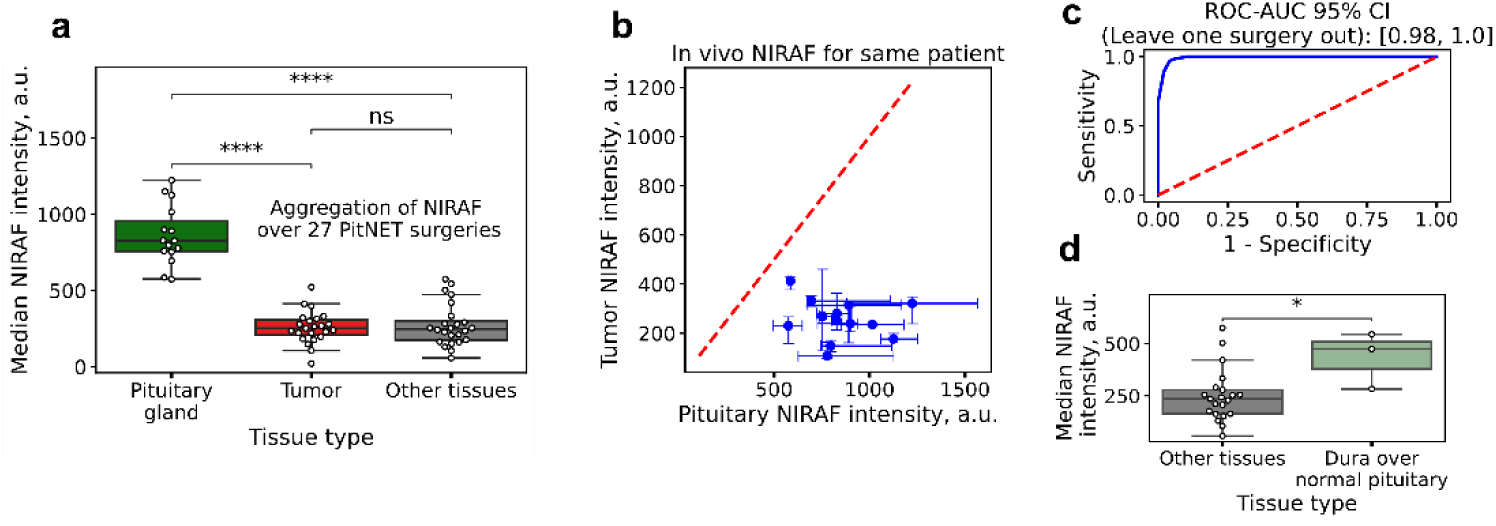
(a) Pooled boxplots of average NIRAF intensity across 27 PitNET surgeries for three tissue classes: pituitary gland, tumor, and other measurement locations. (b) Paired comparison for surgeries with both pituitary and tumor measurements; the dashed line indicates equality. (c) Receiver operating characteristic curve for pituitary versus non-pituitary discrimination (tumor and other measurement locations combined) evaluated using leave-one-surgery-out cross-validation, with AUC and 95% confidence interval shown. (d) Stratification of the other tissues class indicating that the highest intensities are primarily contributed by measurements from the dura overlying normal pituitary.

To evaluate within-patient consistency, we analyzed the subset of surgeries with paired measurements obtained from both normal pituitary and PitNET tissue. In the paired scatter plot, all points lie below the line of equality, indicating that pituitary NIRAF intensity was higher than tumor NIRAF intensity for every individual patient, with no cases showing the opposite trend (Fig. 6b, paired t-test, *p* < 10^-3^). We next quantified discriminative performance using receiver operating characteristic analysis for classification of pituitary versus non-pituitary sites (tumor and other measurement locations combined) with leave-one-surgery-out cross-validation (Fig. 6c). In this procedure, each surgery is iteratively held out for testing, while the decision threshold is derived from the remaining surgeries; aggregating predictions across all held-out folds yields an AUC of 0.99 with a 95% confidence interval of 0.98 to 1.00, demonstrating near-perfect separability between pituitary and non-pituitary measurements. Finally, stratifying the heterogeneous other tissues class showed that the highest intensity values predominantly originated from measurements of the dura overlying normal pituitary, whereas the remaining other sites clustered at lower intensities (Fig. 6d), explaining the upper tail and partial overlap observed in Fig. 6a.

## 4. Discussion

The practical goal of this study was to evaluate whether NIRAF, quantified intraoperatively, can provide an additional optical contrast during endoscopic endonasal transsphenoidal pituitary surgery for distinguishing normal pituitary gland from tumor and surrounding tissues for supporting gland preservation during resection. Across 27 surgeries, pituitary measurements exhibited higher NIRAF intensity than tumor and dura measurements (Fig. 6a, b). A representative case (Fig. 5a, c, d) and a continuous-pass experiment (Fig. 5e) illustrate step-like signal changes as the probe traverses different tissue areas.

As the molecular identity of fluorophore(s) underlying pituitary NIRAF in endocrine organs remains unknown, our interpretation is intentionally restricted to microstructural determinants resolved by confocal spectral imaging. *Ex vivo* imaging consistently identified secretory granule ROIs as the dominant microscopic compartment with the most long-wavelength emission among the major tissue constituents. Under 640-nm excitation, the granule fluorescence spectral shape was largely conserved across diagnostic groups, whereas group differences were expressed primarily in the amplitude of integral granule emission and area. Hence, intraoperative NIRAF contrast is driven mainly by quantitative differences in the contribution of granule-rich microdomains, including ROI area fraction, rather than by a qualitative shift in the intrinsic fluorophores spectral profile.

In addition, collagen-related fluorescence under 405 nm excitation was more prominent in normal pituitary than in tumor tissue (Fig. 3c). Hence, we consider that adding the second fluorescence channel in the setup may allow for additional contrast between pituitary, PitNETs and surrounding tissues.

The continuous-pass trace (Fig. 5e) demonstrated that the NIRAF signal can change over short timescales as the fiber probe traverses adjacent structures within the confined surgical corridor, producing plateau-like segments corresponding to visually identified tissue types. This observation is consistent with the feasibility of using NIRAF as an intraoperative adjunct contrast readout under routine workflow constraints, while interpretation relies on contact geometry and mixed tissue composition at tissue interfaces. Overall, when aggregated across 27 surgeries, NIRAF demonstrated a near-perfect ability to discriminate healthy pituitary tissue from tumor tissue, with a systematically high difference in NIRAF values between pituitary and tumor tissues across all patients (Fig. 6a). Importantly, the separation was patient-invariant in paired sampling (Fig. 6b), supporting the use of NIRAF as a relative, within-case safeguard signal despite inter-surgery variability. Consistently, leave-one-surgery-out validation (Fig. 6c) indicates that the contrast is transferable across surgeries rather than driven by a few favorable cases, yielding an AUC of 0.99 with a 95% confidence interval of 0.98 to 1.00.

A practical feature of the combined cohort analysis was partial overlap between tissue classes and occasional outliers, including sporadically elevated values in classes unrelated to the pituitary gland. Based on synchronized review of the endoscopic video and intraoperative labeling, we associated these events with the following issues. First, some elevated values occurred at sites labeled intraoperatively as dura over the pituitary gland (Fig. 6d). Elevated readings at dura-over-gland sites may reflect proximity effects, where the collected signal partially originates from the underlying glandular tissue through a thin dural layer. This scenario is not clinically ambiguous because dura is visually distinct from the gland under white-light endoscopy, and such optical outliers do not interfere with surgical identification. In the context of gland preservation, these points can be interpreted as measurements unrelated to the gland, even when signal values are elevated. Second, other elevated non-pituitary points occurred when the probe contacted an inhomogeneous surface or an interface, for example at the junction between the pituitary gland and neighboring structures or when pituitary tissue was partially covered with blood. At interfaces (e.g., gland/tumor or gland/dura boundaries), simultaneous measurements of two tissue types are possible, resulting in intermediate (“averaged”) signal levels compared with pure tissue sites. From a practical perspective, such boundary-related changes can be informative because a transition from higher to lower signal during probe movement may help delineating gland boundaries against adjacent tissues during resection. Conversely, low pituitary readings were most observed during episodes of heavy bleeding or when a blood film covered the tissue surface.

In addition to the gland, tumor, and dura, the surgical corridor may include other visually identifiable structures that surgeons may designate as the diaphragm or cavernous sinus, depending on the stage of surgery and anatomical distortion. In our dataset, these areas were grouped into the other tissues category during sampling.

The present work focuses on distinguishing normal pituitary gland from non-pituitary tissues using a single signal, NIRAF intensity. A future direction is to extend optical readouts to better separate low-signal tissue classes, in particular tumor versus adjacent non-pituitary structures. This feature can be especially useful in difficult situations, such as invasive growth, when precise detection of tumor boundaries may contribute to a more complete resection while preserving the normal gland.

## 5. Conclusion

We showed that label-free NIRAF (excitation = 650 nm/emission = 805 nm) is a promising intraoperative modality for distinguishing the normal pituitary gland from tumor and adjacent tissues during transsphenoidal surgery. *Ex vivo* confocal imaging identified secretory granules with a broad excitation spectrum (at least in the 405-640 nm range) is likely a microstructural correlate of NIRAF. In parallel, the molecular origin of granule-associated autofluorescence is being investigated. Identifying the fluorophore or fluorophores responsible for long-wavelength emission is an important next step that can strengthen the mechanistic interpretation and guide further optimization of the intraoperative optical approach.

## Data Availability

The datasets generated and analyzed during the current study are not publicly available due to ethical and legal restrictions related to patient confidentiality. All summary data necessary to evaluate the conclusions are presented in the manuscript.

